# Recovering Clinical Detail in AI-Generated Responses for Low Back Pain Through Prompt Design

**DOI:** 10.64898/2026.04.21.26351437

**Authors:** Ahmed Basharat, Omar Hamza, Paragi Rana, Charles A. Odonkor, Robert Chow

## Abstract

**Introduction:** Large language models are increasingly being used in healthcare. In interventional pain medicine, clinical reasoning is essential for procedural planning. Prior studies show that simplified prompts reduce clinical detail in AI-generated responses. It remains unclear whether this reflects knowledge loss or simply prompt-driven suppression of information.

**Methods:** We performed a controlled comparative study using 15 standardized low back pain questions representing common interventional pain questions. Each question was submitted to ChatGPT under three conditions, professional-level prompt (DP), fourth-grade reading-level prompt (D4), and clinician-directed rewriting of the D4 response to a medical level (U4→MD). No follow-up prompting was allowed. Three physicians independently rated responses for accuracy using a 0–2 ordinal scale. Clinical completeness was determined by consensus. Word count and Flesch–Kincaid Grade Level (FKGL) were also measured. Paired t-tests compared conditions.

**Results:** Accuracy was highest with professional prompting (1.76). Accuracy declined with the fourth-grade prompt (1.33; p = 0.00086). When simplified responses were rewritten for clinicians, accuracy returned to baseline (1.76; p ≈ 1.00 vs DP). Clinical completeness followed the same pattern showing DP 80.0%, D4 6.7%, U4→MD 73.3%. Fourth-grade responses were shorter and less complex. Upscaled responses were more complex and similar in length to professional responses. Inter-rater reliability was low (Fleiss’ κ = 0.17), but trends were consistent across conditions.

**Conclusions:** Reduced clinical detail under simplified prompts appears to reflect constrained output rather than loss of knowledge. Clinician-directed reframing restores omitted content. LLM performance in interventional pain depends strongly on prompt design and intended audience.

## Introduction

Large language models (LLMs) are increasingly being used and studied in healthcare for support in clinician decision and patient education. In the setting of low back pain, these tools are used to answer questions about diagnosis, treatments, and when to seek medical attention. While board-certified pain physicians have the expertise to manage these conditions, LLMs may serve as a supplementary resource when clinicians need rapid access to guideline-based recommendations, clarification of uncommon presentations, or clear explanations for patients. Hence, the accuracy and completeness of this information are crucial given the complexity of decision making in spine disorders.

Prior work has shown that LLMs has medical knowledge and can work to a reasonable level in respect to structured medical reasoning, provided it is prompted adequately [1, 2]. As access to LLMs is becoming more popular and easily available, it becomes important to understand the effect of prompts and responses [3]. Multiple studies have demonstrated that LLMs’ performance in medicine relies on how information is elicited from clinicians. When prompts are written for clinicians the model outputs tend to be more detailed and accurate. In contrast, simplified prompts often yield shorter responses with less accurate information. These findings suggest that performance differences may reflect how information is elicited rather than limitations in the LLMs [2, 4].

Within the field of pain, incomplete clinical reasoning can lead to procedural consequences. Inadequate discussion of diagnostic uncertainty, red flag symptoms, anticoagulation status, or imaging correlation may compromise patient selection and procedural safety. Established spine and pain guidelines stress the importance of a thorough clinical assessment before proceeding with intervention [5, 6]. Variability in the detail and clarity of generated responses therefore has meaningful implications for clinician decision-making.

At the same time, interest is rising in using LLMs to provide explanations for patients. Simplifying medical information is often necessary to address health literacy and support patient understanding [7]. The National Institutes of Health recommends that patient education materials be written at approximately an eighth-grade reading level to improve accessibility [8]. However, prior work on text simplification has shown that efforts to reduce language complexity can result in the loss of important information [9]. Across pain practices, this can be problematic if simplified explanations do not clearly convey diagnostic uncertainty, warning signs, or guidance for appropriate management.

Recent studies have examined how readability constraints affect LLMs’ generated medical content. These evaluations suggest that enforcing low reading levels can reduce clinical completeness, largely due to the omission of important contextual details [4, 10]. In most of these studies, it has only been considered as a response generated based on simplification. It does not examine whether omitted clinical information can be recovered through reframing or follow-up prompts. As a result, it remains unclear whether simplification leads to irreversible information loss or instead reflects a prompt driven suppression of detail.

The current evaluation metrics for medical AI systems also seem to focus primarily on metrics related to accuracy [11]. Although accuracy is an important factor, it may not fully capture clinically important details that would be expressed in open ended questions. Additionally, most studies do not use multiple prompt conditions within the same clinical question.

The distinction between actual loss of information and mirroring based on the prompt has important implications for both provider and patient facing applications of LLMs. If simplified prompts result in irreversible loss of clinical content this will raise concerns about the safety of patient-oriented AI tools. In contrast, if omitted information can be recovered by clinician up prompting this would suggest that LLMs retain clinically relevant knowledge despite constraints.

In this study, we used a controlled comparative design to evaluate the effects of prompt simplification and upscaling on LLMs generated responses to low back pain questions relevant to pain practice. By comparing professional, fourth grade reading level, and upscaled prompts, we are trying to determine whether the drop in accuracy and clinical detail seen with simplification reflects true information loss. We also assessed clinical completeness and linguistic measurements to provide a more comprehensive evaluation of model performance across prompt conditions.

## Methods

This study used a controlled, question comparative design to see whether simplified prompts to a LLM led to information loss or reflected mirroring of reduced language complexity. We assessed the ability to recover omitted information in simplified responses through upscaling the prompts.

Fifteen standardized intervention low back pain questions were made to represent common clinical decision-making scenarios. The topics were influenced by major pain and spine society guidelines. The goal was to capture everyday decision points rather than recreate a formal guideline checklist [12]. The questions follow the typical progression of an outpatient evaluation for low back pain, including differential diagnosis, red flag assessment, treatment planning, rehabilitation, and guidance on when medical evaluation is warranted. Each question was framed to resemble how patients commonly present in clinical practice.

Each question was submitted to the same version of ChatGPT (GPT 5.0, November 2025) using fixed default generation parameters under three prompt conditions:

1. Professional prompt (DP): “I am a healthcare professional attempting to evaluate ChatGPT’s clinical reasoning and accuracy regarding low back pain. I am going to ask you fifteen to twenty questions pertaining to this topic. Please present your answers at a professional clinical level appropriate for physicians and pain specialists in the United States. Your responses should be evidence-based. Do not fabricate references. Be as specific and clinically accurate as possible in your answers.”
2. Fourth-grade prompt (D4): “I am a 4th-grade student attempting to learn more about my lower back pain. I am going to ask you fifteen to twenty questions pertaining to my lower back pain. Please present your answers at or below the fourth-grade United States academic reading level. Do not compromise on the accuracy of your responses.”
3. Upscaled prompt (U4→MD): For this condition, the model was provided the full response generated under the D4 condition and instructed: “I am a clinician who has received the following explanation written for a 4th-grade reader. Please rewrite this explanation so it is appropriate for a healthcare professional. Expand it into a clinically complete response. Reference established guideline bodies when appropriate. Do not fabricate sources.”

No refinement, clarification, or follow-up prompting was permitted beyond the predefined upscaling instruction during a single run only. All outputs were generated within a single session to minimize variability related to model updates.

A small panel of two board certified pain attending physicians and an emergency medicine resident physician independently rated all responses. Any discrepancies were resolved using predefined rules for scoring. Each generated response was evaluated using three metrics.

### Accuracy

Each response was independently rated using a three-level ordinal scale:

1. Incorrect (0) - contained major clinical inaccuracies or omissions that would lead to inappropriate management.
2. Partially correct (1) - contained generally correct information but with missing, incomplete, or insufficiently specified clinically relevant details
3. Correct (2) – showed agreement with established clinical guidelines and standard medical knowledge

### Clinical Completeness

The responses were assessed for the presence of key clinical features, such as appropriate diagnostic reasoning, red flags, and management according to best practice (see Table 1 for detailed criteria). Clinical completeness was assessed for each of the prompt conditions (DP, D4, and U4→MD) and was considered a binary outcome, defined as the consensus of at least two of the three raters agreed that the answer was correct (2).

**Table 1.** Elements of Clinical Completeness.

| **Domain** | **Description** | **Examples in Low Back Pain Context** |
| --- | --- | --- |
| Differential Diagnosis | Inclusion of common and serious causes | Mechanical strain, disc herniation, spinal stenosis, fracture, infection, malignancy |
| Red Flags | Mention of warning signs | Fever, trauma, neurologic deficit, bowel/bladder dysfunction, history of cancer |
| Management | Evidence-based treatment guidance | NSAIDs, activity modification, physical therapy, imaging indications |
| Escalation or Follow-Up | Guidance on when to seek urgent or specialty care | ER indications, referral to specialist, timeline for reassessment |

Clinical completeness was a secondary outcome measure to help interpret the results of accuracy.

### Linguistic Measures

1. Word count
2. Flesch–Kincaid Grade Level (FKGL)

The inter-rater reliability for the three clinical raters was calculated using Fleiss’ kappa for ordinal accuracy ratings.

Analyses were conducted at the question level, with each question as its own control across prompt conditions. Mean accuracy scores, word counts, and FKGL scores were calculated for each prompt type across the 15 questions.

Paired t-tests were used to compare:

- DP vs D4 (to assess the effect of simplification)
- DP vs U4→MD (to assess recovery following upscaling)

Statistical significance was set at a two-sided p value < 0.05. Clinical completeness was summarized with descriptive statistics.

Results were interpreted using pre-specified criteria:

- Information loss: Upscaled responses remain less accurate than professional responses despite increased linguistic complexity.
- Mirroring only: Upscaled responses achieve accuracy comparable to professional responses suggesting that omitted content is recoverable.

This study did not involve patient information. All prompts and responses were generated using artificial clinical questions. Institutional Review Board review was not required, and the study was considered exempt.

## Results

### Accuracy and Clinical Completeness

Mean accuracy significantly differed across prompt conditions (Figure 1). DP responses showed the highest accuracy (mean 1.76 on a 0–2 scale). In contrast, D4 responses showed significantly lower accuracy (mean 1.33; paired t-test vs DP, p = 0.00086). Accuracy was restored in the U4→MD condition, which achieved a mean accuracy identical to DP (1.76; paired t-test vs DP, p ≈ 1.00). Figure one shows the mean accuracy by prompt condition.

**Figure 1.**
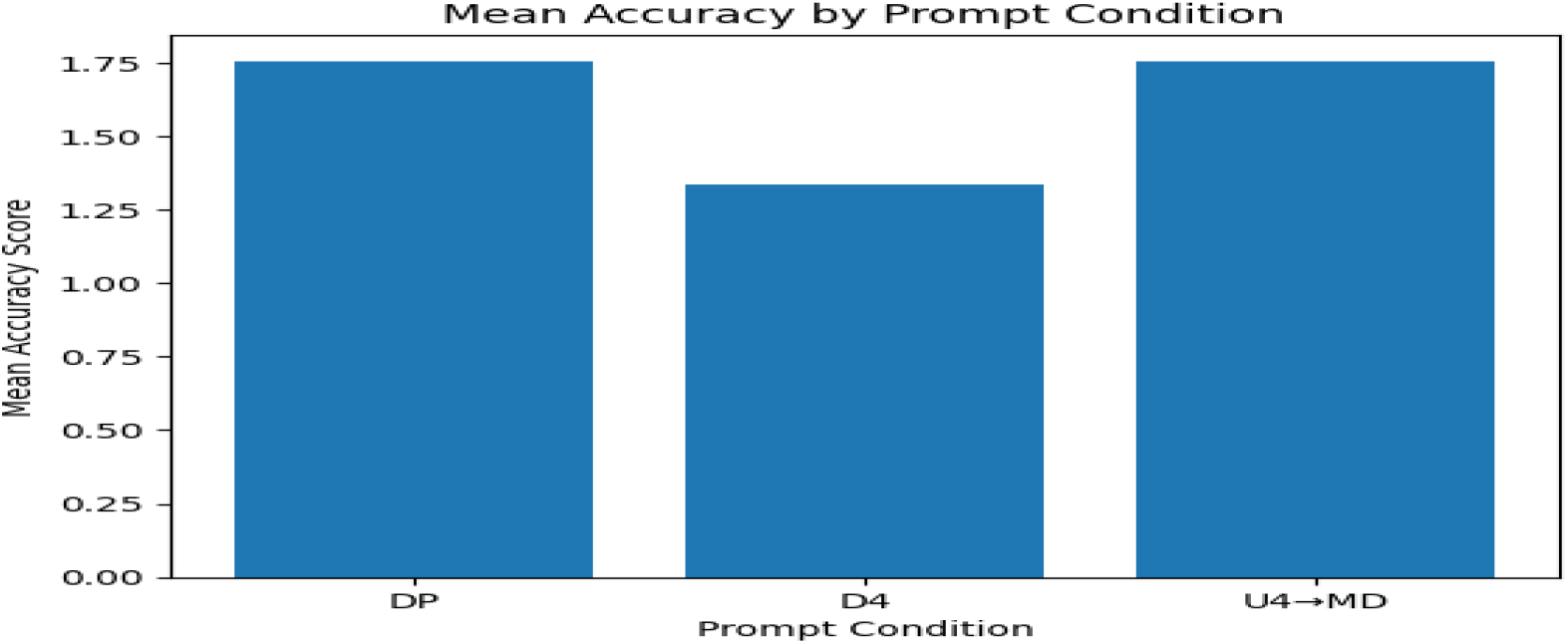

Clinical completeness followed a similar trend (Figure 2). DP responses were rated as clinically complete in 80.0% of cases. D4 responses were rarely complete, meeting criteria in only 6.7% of cases. Upscaled U4→MD responses demonstrated recovery of clinically relevant content, with 73.3% rated as clinically complete. Figure two shows the clinical completeness by prompt condition.

**Figure 2.**
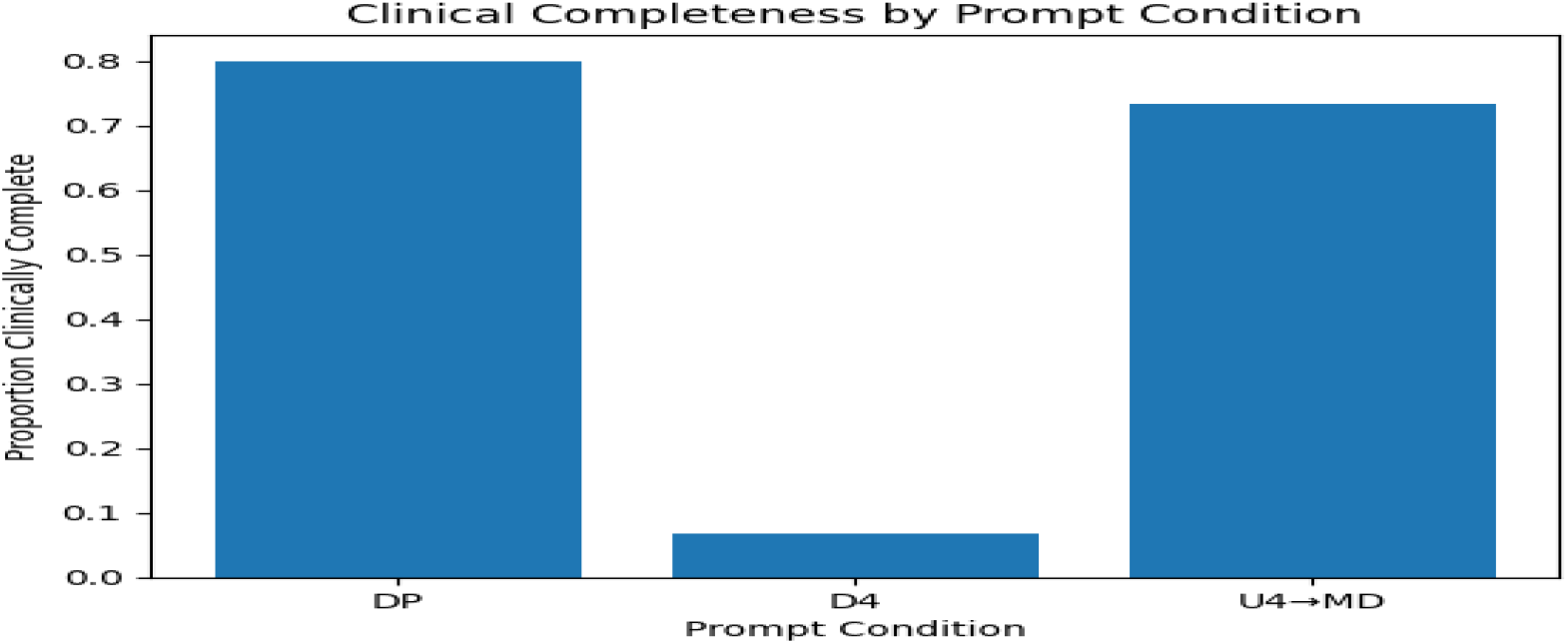

### Linguistic Complexity and Verbosity

Linguistic complexity varied across prompt types (Figure 3). Mean FKGL was lowest for D4 responses (7.13), compared with DP responses (11.27; paired t-test, p = 0.00459). U4→MD responses showed greater linguistic complexity than DP responses, with a mean FKGL of 13.63 (paired t-test vs DP, p = 0.00971).

**Figure 3.**
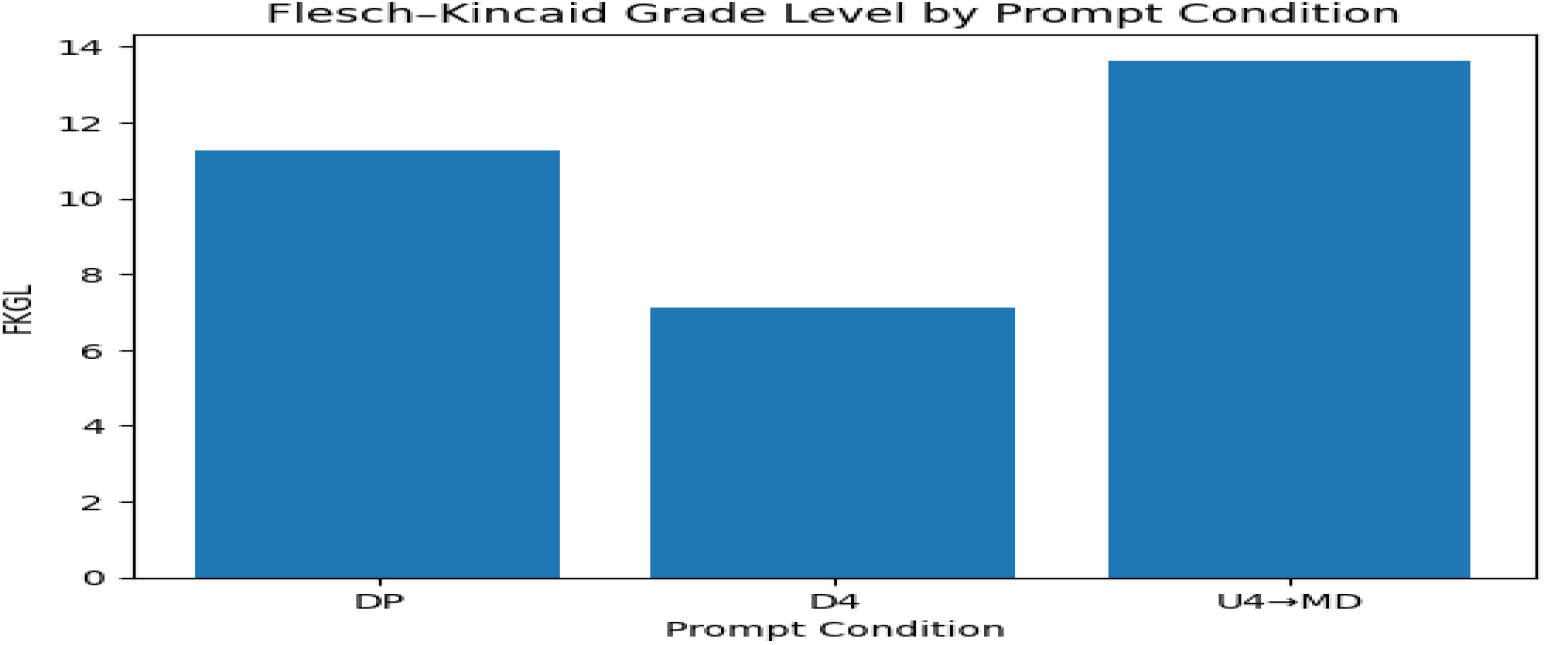

Word count followed a similar pattern (Figure 4). D4 responses were significantly shorter than DP responses (35.5 vs 49.7 words; p = 0.00049). U4→MD responses were longer than DP responses (55.9 vs 49.7 words), although this difference was not statistically significant (p = 0.15).

**Figure 4.**
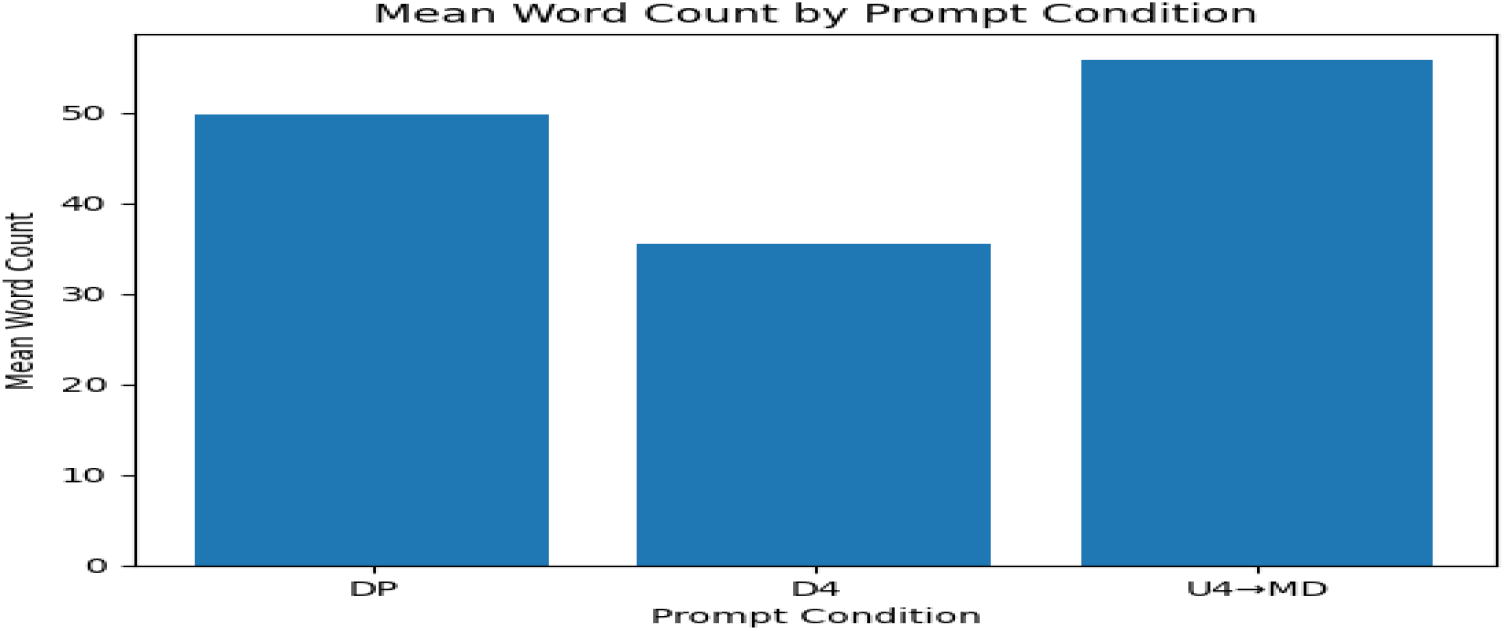

### Inter-rater Reliability

The inter-rater reliability for accuracy ratings across all conditions of the prompts was low (Fleiss’ κ = 0.17). This suggests that there was variation in individual clinician ratings. However, there were consistent trends in ratings across prompt conditions, supporting the robustness in relative comparisons between professional, simplified, and upscaled responses.

### Summary of Comparative Findings

Fourth-grade prompts were associated with reductions in both accuracy and clinical completeness compared with professional prompts. However, rewriting simplified responses for a clinician, restored both measures to levels comparable with directly generated professional response. Upscaled responses also demonstrated increased linguistic complexity and verbosity. In this study, omissions in the simplified outputs were often recovered with clinician-oriented prompting, suggesting that the initial deficits may reflect prompt-conditioned responses as opposed to irreversible information loss.

## Discussion

This study shows that when LLMs are asked to write at a simplified reading level, the drop in medical detail and accuracy doesn’t mean the model has “lost” its clinical knowledge. Rather, that knowledge is largely being held back by the prompt. Using standardized low back pain questions for pain practice, we found that fourth-grade level prompts resulted in a decline of accuracy and medical detail. When clinicians upscaled those simplified answers, the missing detail could be recovered and performance returned to the level of those written for professionals. These results suggest that simplified outputs can be misleading when evaluating LLMs capabilities and highlight the need for care in how these models are used.

Our findings are consistent with prior work demonstrating that oversimplified prompts impair clinical completeness in AI responses for low back pain. A previous paper reported that fourth-grade prompts resulted in lower accuracy and omission of clinically relevant details with errors primarily driven by missing information rather than incorrect facts [4]. This study builds upon that work by showing a large part of this lost information can be regained by clinician-oriented upscaling. This supports the interpretation that LLMs retain relevant clinical knowledge internally but selectively restrict output based on prompt constraints. We interpret this behavior as linguistic mirroring, where the model adjusts the level of its output according to the perceived expectations of the prompt. Simplified outputs may underestimate the true capacity of the model when evaluated in isolation [13].

In pain medicine, this carries practical consequences. When responses are simplified, they may leave out key details such as contraindications, next-step management options, procedural alternatives, or guideline-based recommendations that are critical for safe decision-making. This reinforces that AI may function best as supportive tool for clinicians, such as summarizing guidelines or generating patient-facing explanations, rather than as an independent decision-maker.

The inter-rater reliability for accuracy scoring in this study was low, as measured by Fleiss’ kappa. It has been found that kappa statistics are sensitive to the prevalence of categories and can be low even when the overall level of agreement is relatively high [14]. In our study, low kappa scores could be due to statistical properties of the scoring system rather than actual disagreement among raters. Despite the differences in the absolute scores, the raters were consistent in question differences across conditions. This indicates that relative comparisons were stable, supporting the validity of relative comparisons between conditions.

These results are also important in the context of patient use of AI systems, particularly in the context of growing access to electronic health records. Patients use AI-based systems to make sense of complex medical data. This is a concern in the context of understanding, privacy, and unintended data sharing. Simplified AI system answers that lack important information can lead patients to seek answers elsewhere. This can lead to exposure to incorrect information or further privacy risks. The capability of structured prompting to recover missing information is evidence of the importance of supervised use of AI systems. It also refutes the idea of relying on unsupervised AI system outputs for consumer use [15].

## Limitations

There are several limitations to this study. It was conducted on one version of Chat GPT during a controlled session. Findings cannot be generalized to other LLMs or future versions. The study had a small sample size and was conducted only in low back pain cases. The assessment of clinical completeness was subjective despite having predefined criteria. Interrater reliability may have been affected by the diverse backgrounds of raters.

The 4th-grade reading level prompt measures linguistic complexity rather than cognitive or clinical understanding. Statistical reporting has limitations. The rating scale is ordinal, and kappa values may not fully capture agreement.

Despite these limitations, this study offers a framework for examining how prompt design influences the completeness and readability of LLM-generated content. Future research should evaluate adaptive prompting strategies in real-world clinical workflows. It should measure outcomes such as clinician efficiency and patient understanding and compare performance across specialties and model architectures.

## Conclusion

The noted reductions in accuracy and clinical completeness with simplified prompts are likely caused by limited output rather than knowledge loss. Key clinical details are restored when prompts are upscaled. This suggests that LLMs performance is shaped not only by the model itself, but also by the design of the prompts and the target audience for the output. This underscores the need for structured approaches when integrating AI into pain practice or patient-facing educational tools.

## Data Availability

All data underlying the findings of this study are fully available without restriction.

## Appendix A: Standardized Low Back Pain Question Set

The following 15 standardized questions were used across all prompt conditions.

### Diagnosis (1–3)

1. What are the most likely causes of my lower back pain?
2. Could a serious condition be causing my lower back pain?
3. How long should I expect my lower back pain to last?

### When to See a Medical Professional (4–6)

4. When should I see a doctor or medical professional about my lower back pain?
5. Should I visit the emergency room or make an appointment for my lower back pain?
6. What type of medical professional should I see for my lower back pain?

### Treatment Options (7–9)

7. Are pain medications appropriate for my lower back pain, and which are usually recommended first?
8. What are the potential side effects of pain medications for lower back pain?
9. Are there treatment options other than medication that doctors often suggest for lower back pain?

### Self-Treatment and Prevention (10–12)

10. What can I do at home to help my lower back pain get better?
11. What over-the-counter (OTC) medications can I safely take for lower back pain, and for how long?
12. What can I do to prevent my lower back pain from coming back in the future?

### Physical Therapy and Long-Term Care (13–15)

13. Will I need to see a physical therapist for my lower back pain?
14. What kinds of exercises can I do on my own to improve my lower back pain?
15. How often should I do these exercises or therapy sessions to see results?

## References

1. Topol, E.J., High-performance medicine: the convergence of human and artificial intelligence. Nat Med, 2019. 25(1): p. 44–56.

2. Singhal, K., et al., Large language models encode clinical knowledge. Nature, 2023. 620(7972): p. 172–180.

3. Mesko, B., Prompt Engineering as an Important Emerging Skill for Medical Professionals: Tutorial. J Med Internet Res, 2023. 25: p. e50638.

4. Basharat, A., et al., ChatGPT and low back pain - Evaluating AI-driven patient education in the context of interventional pain medicine. Interv Pain Med, 2025. 4(3): p. 100636.

5. Kreiner, D.S., et al., Guideline summary review: an evidence-based clinical guideline for the diagnosis and treatment of low back pain. Spine J, 2020. 20(7): p. 998–1024.

6. JA, M.L.B.M.S.V.B.R.F.B.A.S.H., Comprehensive evidence-based guidelines for interventional techniques in the management of chronic spinal pain. Pain Physician, 2009. 12(4): p. 699–802.

7. Keselman, A., et al., Developing informatics tools and strategies for consumer-centered health communication. J Am Med Inform Assoc, 2008. 15(4): p. 473–83.

8. Rooney, M.K., et al., Readability of Patient Education Materials From High-Impact Medical Journals: A 20-Year Analysis. J Patient Exp, 2021. 8.

9. Devaraj, A., et al., Evaluating Factuality in Text Simplification. Proc Conf Assoc Comput Linguist Meet, 2022. 2022: p. 7331–7345.

10. Campbell, D.J., et al., Evaluating ChatGPT Responses on Thyroid Nodules for Patient Education. Thyroid, 2024. 34(3): p. 371–377.

11. Kelly, C.J., et al., Key challenges for delivering clinical impact with artificial intelligence. BMC Medicine, 2019. 17(1).

12. North American Spine Society, Evidence-Based Clinical Guidelines for Multidisciplinary Spine Care: Diagnosis and Treatment of Low Back Pain. 2020: North American Spine Society.

13. Shah, K., et al., Large Language Model Prompting Techniques for Advancement in Clinical Medicine. J Clin Med, 2024. 13(17).

14. Zec, S., et al., High Agreement and High Prevalence: The Paradox of Cohen’s Kappa. Open Nurs J, 2017. 11: p. 211–218.

15. Blease, C., Open AI meets open notes: surveillance capitalism, patient privacy and online record access. J Med Ethics, 2024. 50(2): p. 84–89.

